# Fingerprint method applied to data from a phase III clinical trial

**DOI:** 10.1101/2024.06.25.24309472

**Authors:** Lars Edenbrandt

## Abstract

Researchers in the RECOMIA network have been developing AI tools for the automated analysis of PET/CT studies in lymphoma patients. To enhance these AI tools, the CALGB 50303 dataset from The Cancer Imaging Archive was identified for inclusion in their project. Ensuring the quality of databases used for AI training is crucial, and one quality control (QC) measure involves the AI-based Fingerprint method to verify correct de-identification of clinical trial images. The study applied the Fingerprint method to PET/CT studies from 130 patients, successfully detecting an incorrectly de-identified study and identifying its correct trial identification number. This demonstrates the feasibility of using AI for QC in clinical trials. AI-based methods offer significant opportunities for enhancing QC, providing automated, consistent, and objective analyses that reduce the workload on human annotators. Integrating AI into QC processes promises to improve accuracy, consistency, and efficiency, thereby enhancing data integrity and the reliability of clinical trial results. This study underscores the importance of further developing AI-based QC methods in clinical trials.

## Introduction

Researchers in the RECOMIA (www.recomia.org) network have been developing and validating AI tools for the automated analysis of PET/CT studies in patients with lymphoma for several years [1-3]. These AI methods are data-driven, and there is always an incentive to incorporate new databases into this research. The Cancer Imaging Archive (TCIA) is an open-access database of medical images for cancer research [4]. For a project aimed at improving the AI tool for the automated analysis of PET/CT studies in lymphoma patients, the data collection CALGB 50303 from TCIA was identified [5].

The quality of databases used in AI training is essential for achieving reliable results. Therefore, we have established routines for quality control (QC). One such check involves applying the recently developed AI-based Fingerprint method (SliceVault AB, Malmö, Sweden) to detect whether studies from clinical trials have been correctly de-identified [6]. The name and identification number of each subject are manually replaced with a trial identification number, and errors in this process can lead to false results. Before using the PET/CT data in the lymphoma project, the Fingerprint method was applied to these studies.

## Methods

The CALGB 50303 data collection was selected and downloaded from The Cancer Imaging Archive (TCIA) [5]. This collection contains data from the National Cancer Institute Clinical Trial NCT00118209, “Rituximab and Combination Chemotherapy in Treating Patients With Diffuse Large B-Cell Non-Hodgkin’s Lymphoma.” This randomized phase III trial studied the effectiveness of rituximab in combination with two different chemotherapy regimens for treating patients with diffuse large B-cell lymphoma. PET/CT studies from a total of 130 patients were downloaded, which included 119 baseline studies and 219 first and second follow-up studies. From these 338 studies, 286 pairs of studies from the same patient were selected.

The first step of the Fingerprint method (SliceVault AB, Malmö, Sweden) involves segmenting the left and right hip bones in the CT images using the AI-based Organ Finder method [7]. The corresponding hip bones from two CT studies are then aligned, and a measure of similarity, known as the anatomical match, is calculated. If the anatomical match is less than 80% for both the left and right hip bones, the two CT studies are classified as being from different subjects [6].

## Results

The anatomical match of the left and right hip bones for the 286 pairs of CT studies is presented in Figure 1. The two data points with an anatomical match around 30% correspond to a patient with three CT studies. One of these studies, when compared to the other two, results in anatomical match values indicating that it was not obtained from the same patient as the other two studies. The comparison between the other two studies results in an anatomical match of 97% for both left and right hip bones, confirming they were obtained from the same patient.

**Figure 1:**
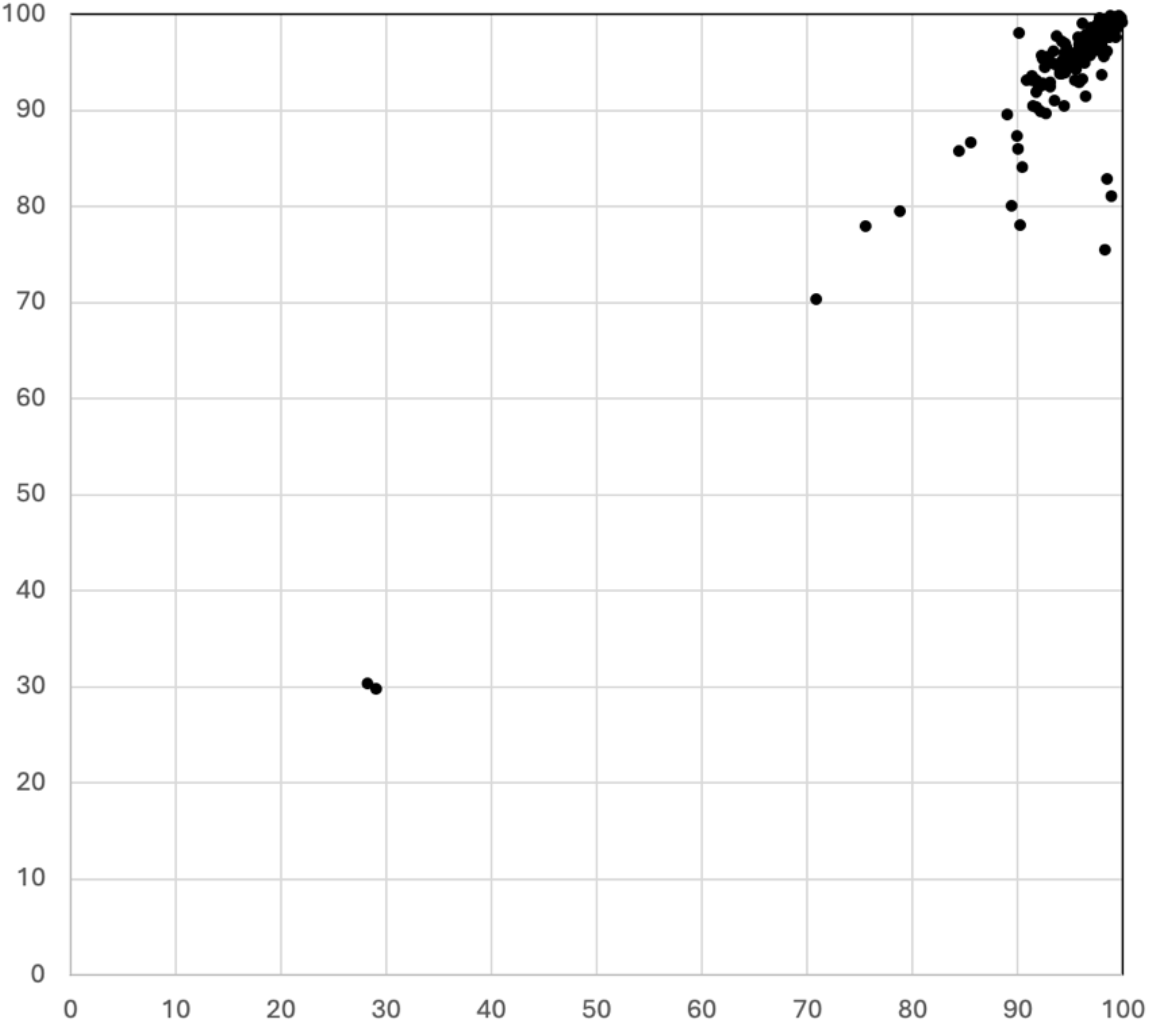
Anatomical match for the left (X axis) and right (Y axis) sided hip bone from pairs of CT studies from the same patient (n=286).

For comparison, the anatomical match for the same number of pairs of CT studies from different patients is presented in Figure 2. In this plot, an anatomical match of about 30% is typical for a pair of CT studies from different patients.

**Figure 2:**
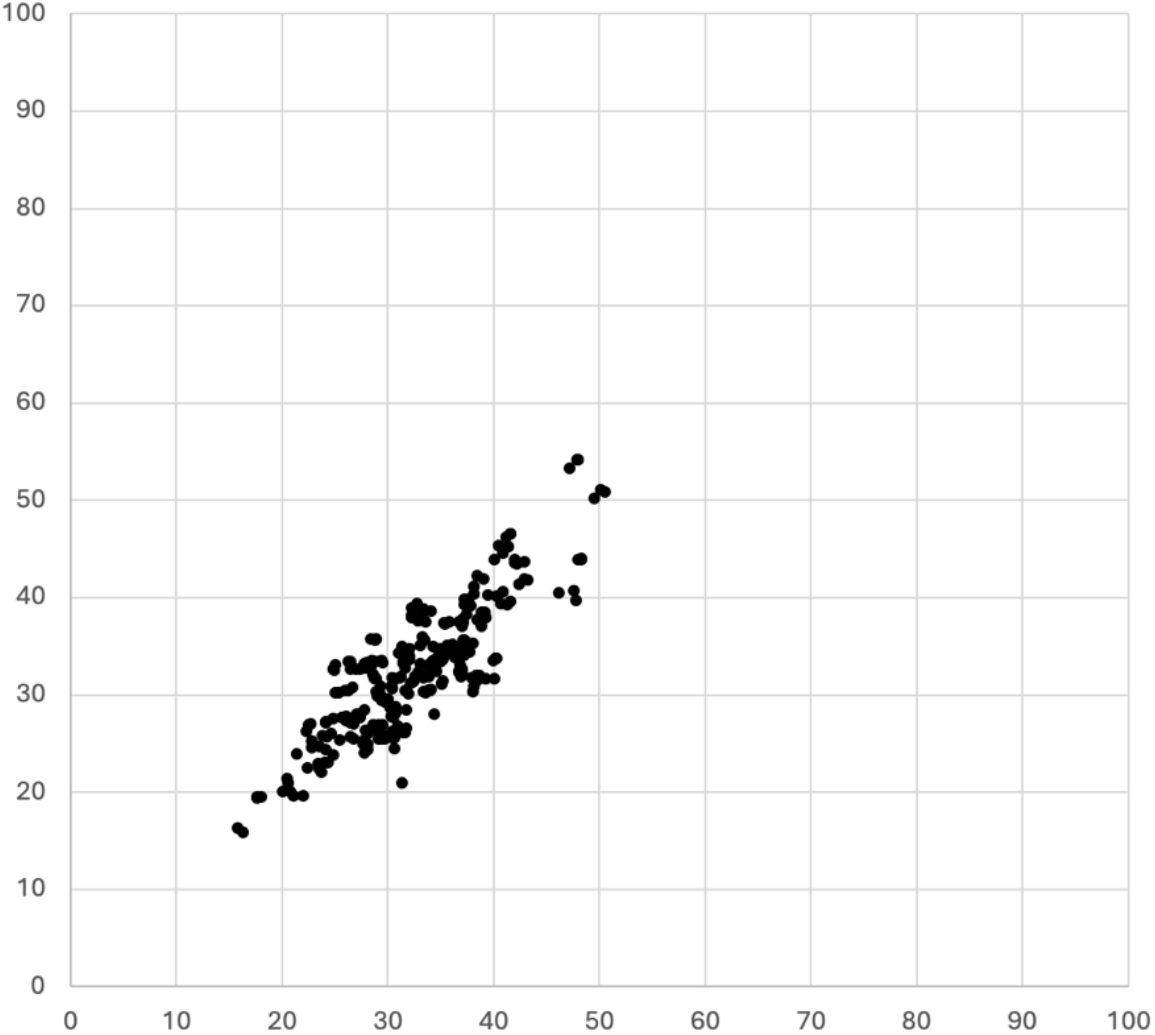
Anatomical match for the left (X axis) and right (Y axis) sided hip bone from pairs of CT studies from different patients (n=286).

One CT study with a mislabeled trial identification number was detected, as shown in Figure 1. In the next step, if this study matched any of the CT studies from the other 129 patients was investigated. One CT study from each of these patients was selected and compared to the mislabeled study using the Fingerprint method. The results are shown in Figure 3. One pair of CT studies showed an anatomical match of 97% for both the left and right hip bones, while all the other pairs displayed low anatomical match values.

**Figure 3:**
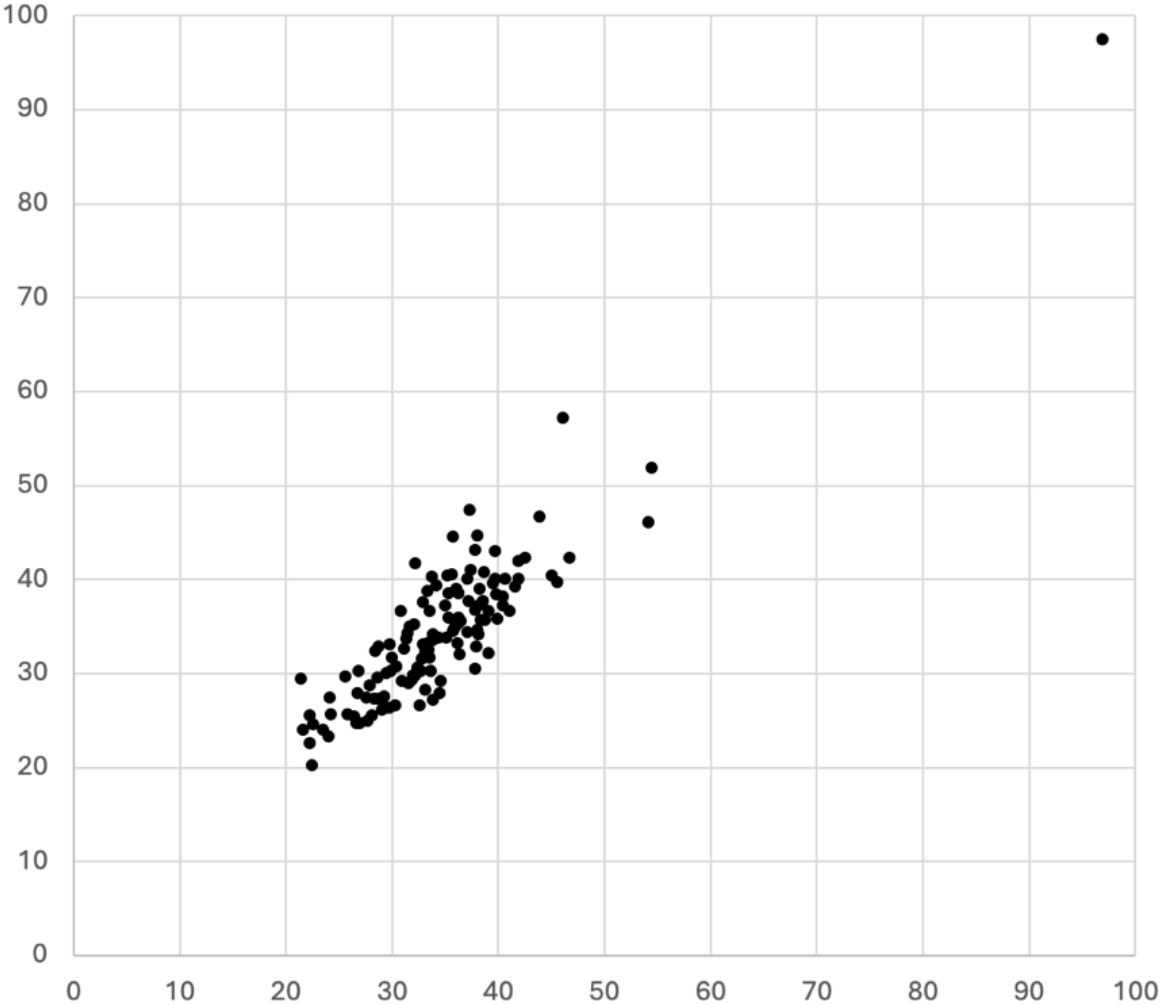
Anatomical match for the left (X axis) and right (Y axis) sided hip bone from pairs of CT studies where one CT from each of 129 patients is compared to the mislabeled study.

The results of the Fingerprint analysis revealed that one of the PET/CT studies was obtained from a different patient than originally indicated. This study was from a female patient, whereas the other two studies were from a male patient. The misidentified study was found to match with a female patient who had another trial identification number. The CT studies from these two patients are shown in Figure 4.

**Figure 4:**
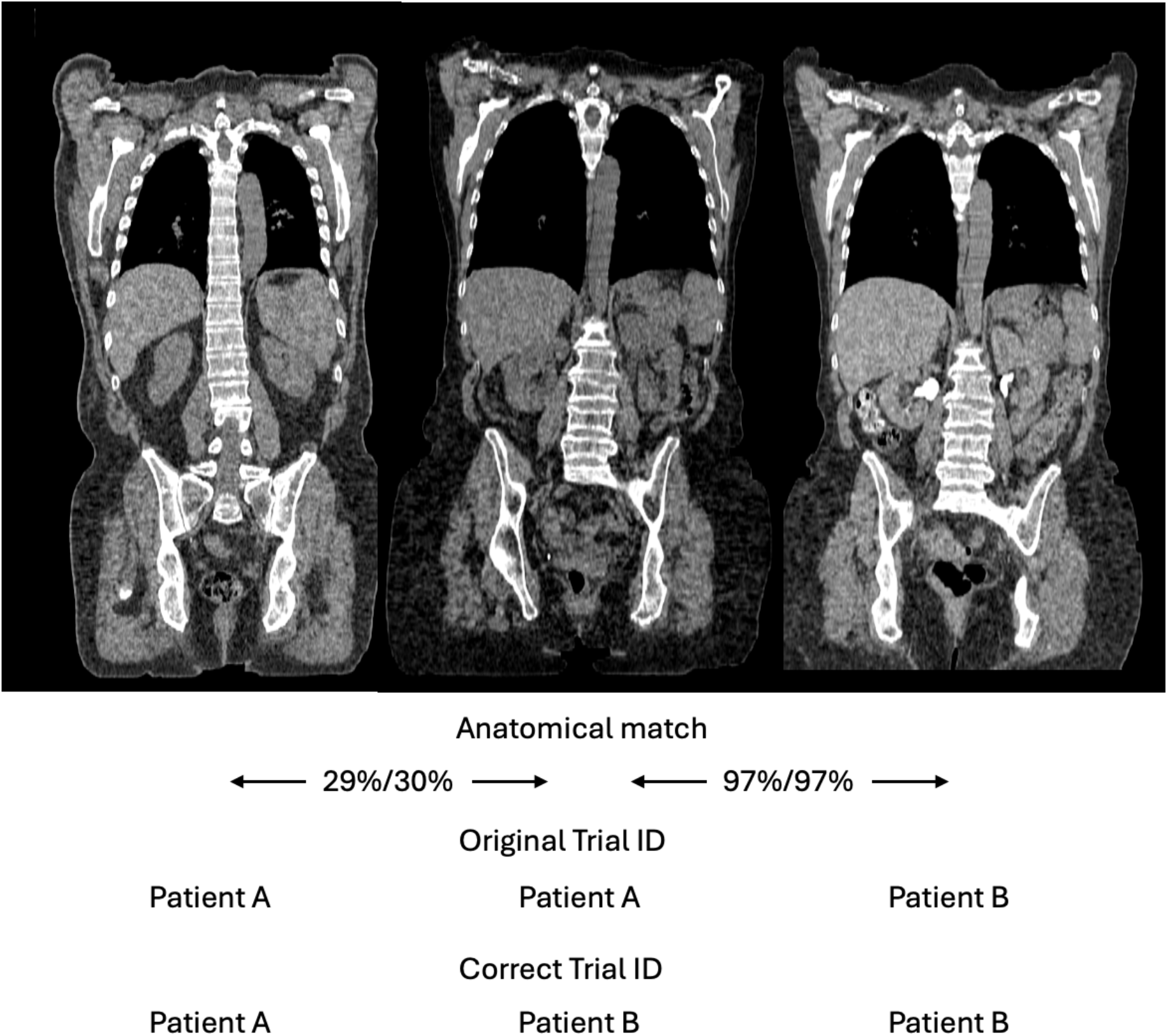
Left and middle CT studies falsely labeled with the same trial identification number were obtained from a male and a female patient. The middle CT study was obtained from a patient with another trial identification number whos CT study is shown to the right.

## Discussion

This study marks the first application of the Fingerprint method to imaging studies from a clinical trial, successfully detecting an incorrectly de-identified PET/CT study and identifying the correct trial identification number for the mislabeled study. The findings demonstrate the feasibility and effectiveness of using an automated AI-based method for QC of images in clinical trials.

De-identification of images in clinical trials involves removing not only names and identification numbers but also information such as patient gender and age. Consequently, patients of different genders or ages can be mislabeled as the same individual. Since only the actual images can be used to detect mislabeling, and image readers may focus more on tumor-related findings than on verifying patient identity across different visits, such errors can go unnoticed. This trial illustrates the value of automated methods like the Fingerprint method in identifying and correcting such issues.

AI offers significant opportunities for enhancing QC in clinical trials. Traditional QC methods often rely on manual processes, which can be time-consuming, prone to human error, and inconsistent. In contrast, AI-based tools can provide automated, consistent, and objective analyses. For example, AI can be used to automatically segment organs and tissues, detect anomalies, and verify the integrity of the data. This reduces the workload on human annotators and quality control personnel, allowing them to focus on more complex tasks that require expert judgment.

In conclusion, the integration of AI into the QC processes of clinical trials holds great promise for improving accuracy, consistency, and efficiency. By leveraging AI tools, researchers can ensure higher standards of data integrity and ultimately enhance the quality and reliability of clinical trial results. This study highlights the importance of continued development and validation of AI-based methods to support robust and reliable QC in clinical trials.

## Data Availability

All data produced in the present study are available upon reasonable request to the authors

https://www.recomia.org/

## Acknowledgment

This manuscript was prepared using data from Dataset CALGB 50303 from the NCTN/NCORP Data Archive of the National Cancer Institute’s (NCI’s) National Clinical Trials Network (NCTN). Data were originally collected from clinical trial “NCT00118209 Rituximab and Combination Chemotherapy in Treating Patients With Diffuse Large B-Cell Non-Hodgkin’s Lymphoma”. All analyses and conclusions in this manuscript are the sole responsibility of the author and do not necessarily reflect the opinions or views of the clinical trial investigators, the NCTN, the NCORP or the NCI.

